# Stroboscopic Light Stimulation in Adults Reporting Depressive Symptoms: Safety, Tolerability, Feasibility, and Active-Comparator Development in a Staged Early-Phase Study

**DOI:** 10.64898/2026.06.17.26355864

**Authors:** Danny Nacker, Luise Kalus, Anil K. Seth, James M. Stone, Robert Chis-Ciure, Gavin Lawson, Jimi Simpson, Josemir W. Sander, Stephen Bremner, Christopher I. Jones, Wendy Wood, Fiona Macpherson, Dirk Proeckl, Engelbert Winkler, David J. Schwartzman

## Abstract

Stroboscopic light stimulation (SLS) is a candidate non-pharmacological intervention that induces transient visual and affective experiences, with potential application in depression. Before efficacy testing, clinical development requires safety, tolerability, and feasibility data. We report a staged, single-site programme in adults reporting depressive symptoms. Study 1 tested 11 SLS parameter sets for safety and tolerability. Study 2 assessed whether an active comparator reduced subjective visual effects while preserving session context. Study 3 randomised 84 participants to four weekly supervised 31-minute sessions of strobe-emphasis or wash-emphasis SLS. In Study 1, 31 participants were analysed; no severe adverse reactions occurred, mean discomfort was low (0.49/10), and the highest session-level upper 80% confidence limit was 1.13/10, well below the prespecified threshold. Study 2 supported within-participant experiential separation between the two SLS configurations in healthy adults. In Study 3, endpoint data were available for 70/84 randomised participants (83.3%): 39/42 assigned to strobe-emphasis and 31/42 assigned to wash-emphasis. Discomfort remained low, with no serious SLS-attributable adverse events. Both configurations produced visual phenomenology in the Study 3 target sample, and the phenomenological separation observed during Study 2 was not reproduced between groups in Study 3. The comparison estimates differences between two active SLS configurations, rather than SLS versus no stimulation. Exploratory depressive-symptom estimates were imprecise, limited by differential endpoint availability, and should inform endpoint selection and subsequent trial design rather than efficacy. Overall, the programme supports further controlled evaluation of supervised SLS while identifying experiential separation, retention, masking, and endpoint selection as priorities for the next trial.

## 1. Introduction

Major Depressive Disorder (MDD) is a common, debilitating, and costly condition, with substantial consequences for individuals, healthcare systems, and society (McCrone et al., 2008; Liu et al., 2020; McDaid et al., 2022). Although antidepressants and psychotherapies can be effective, remission is far from universal, and access remains uneven. Many people do not receive or sustain acceptable care because available treatment pathways can be slow, difficult to access, poorly matched to preferences, or are associated with burdensome side-effects (Arroll et al., 2005; Mendlewicz, 2008; DeRubeis et al., 2008; Cuijpers et al., 2016; Luo et al., 2020; National Institute for Health and Care Excellence [NICE], 2021). There remains a need for brief, scalable, non-pharmacological interventions compatible with supervised clinical delivery.

Stroboscopic light stimulation (SLS) may offer one route. Unlike conventional bright-light therapy, which is usually framed through circadian or seasonal mechanisms, SLS uses controlled rhythmic visual stimulation to induce transient closed-eye perceptual and affective experiences (Mårtensson et al., 2015; Hewitt et al., 2025; Huang et al., 2008; Beauté et al., 2026). Frequency and rhythmic structure shape the intensity and character of these effects, including vivid geometric imagery and other alterations in subjective experience (Amaya et al., 2023; Bartossek et al., 2021; Beauté et al., 2026; Grove et al., 2026; Hewitt et al., 2026b; Schwartzman et al., 2019). For example, Amaya et al. (2023) compared two-minute exposures at 3, 8, 10 and 18 Hz with a 30% duty cycle in healthy adults; geometric and dynamic visual effects were strongest under rhythmic 10-Hz stimulation and weaker under frequency-matched arrhythmic stimulation. The authors reported a lamp output of 5,520 lm at a distance of approximately 1.5 m; this luminous-flux specification is not directly comparable with eye-level illuminance. These findings concern acute phenomenology in screened healthy adults, not antidepressant efficacy. However, systematic evidence defining how stimulation parameters interact remains limited, particularly for longer experience-centred SLS protocols and in populations reporting depressive symptoms. Existing evidence therefore does not identify a single optimal or therapeutically superior SLS parameter set. SLS is therefore clinically interesting not simply as another light-based treatment, but as a controllable, non-pharmacological method for inducing brief experiential change.

The rationale for testing SLS in adults reporting depressive symptoms is that transient perceptual and affective changes may help alleviate depressive symptoms without requiring pharmacological exposure. Alpha-frequency SLS produced antidepressant-like effects in a corticosterone-induced mouse model (Kim et al., 2016), and psychedelic-therapy studies suggest that acute subjective experience can predict later depressive-symptom and wellbeing change (Roseman et al., 2018; Yaden and Griffiths, 2021; Nikolaidis et al., 2023; Klučková et al., 2025; Kettner et al., 2021). These findings provide the rationale for measuring acute experience and later symptom change, but they do not establish that SLS has antidepressant effects or a psychedelic-like mechanism.

Dreamachine public-engagement data provide operational evidence that supervised SLS can be delivered at large scale, with structured screening and supervision, to public audiences. Across a programme involving more than 40,000 members of the public, a subset of respondents reported increases in positive affect and wellbeing after exposure, although these self-reports were not collected under controlled clinical-trial conditions (Dreamachine, n.d.; Beauté et al., 2026). These are not clinical safety or efficacy data.

Together, these uncertainties and the available evidence support a feasibility-first programme to test whether SLS can be delivered safely, tolerably, and consistently in adults reporting depressive symptoms. Symptom outcomes are exploratory signals for later trial design.

We conducted a staged single-site developmental programme to test progression from brief parameter testing to repeated supervised delivery in adults reporting depressive symptoms. Study 1 assessed parameter-level safety and tolerability and identified tolerable parameter settings but did not determine how those settings should be arranged across longer sessions. A reproducible structure inspired by classical sonata organisation was therefore used to organise the sequence rather than as a hypothesised therapeutic mechanism. Study 2 tested whether a wash-emphasis SLS configuration could reduce subjective visual effects while preserving session duration, hardware, music, sequence structure, flicker cues, and overall light exposure. Study 3 then tested a four-session protocol under randomised, allocation-concealed configurations using coded strobe-emphasis and wash-emphasis sequences. Symptom outcomes were exploratory and estimation-focused, consistent with pilot-trial methodology (Whitehead et al., 2016; Teare et al., 2014).

## 2. Methods

This staged single-site programme was conducted at the Sussex Centre for Consciousness Science (SCCS), University of Sussex, using a CE-marked commercial stroboscope (RX1; roXiva Ltd, Sheffield, UK). Programmed light intensity is reported as the RX1 output setting (%) because this permits reproduction on the study device; where available, corresponding physical measurements are reported as illuminance (lux). Illuminance was measured using a SAUTER SO 200K digital light meter positioned at participant eye level 1 m from the stroboscope. At 100% RX1 output, illuminance was approximately 2700 lux at the measurement position. A linear relationship between RX1 output percentage and illuminance was not established, so intermediate settings should not be converted to lux using a fixed multiplier.

Study 1 tested single-visit parameter optimisation in adults reporting depressive symptoms. Study 2 calibrated the control in healthy adults. Study 3 tested repeated SLS delivery in an allocation-concealed randomised feasibility trial using masked sequence codes. Study 1 and Study 3 were recorded under ISRCTN82430224 and ISRCTN13880276, respectively (ISRCTN, 2025a, 2025b). The programme was approved by the University of Sussex Sciences & Technology ethics committee C-REC (ER/LK344/4).

Across all programme stages, SLS was administered individually to seated participants in a closed, soundproofed testing booth. Participants were instructed to keep their eyes closed and were monitored live by a researcher using a camera. Fixed seat and stroboscope positions gave an approximate eye-to-stroboscope distance of 1 m; the exact viewing distance was not recorded for each participant. Study 1 was conducted without musical accompaniment, and during Sessions 1-10 a bell tone marked the 30-s parameter transitions. Study 2 and Study 3 used musical accompaniment. Additional audio specifications are provided in the Supplementary Material.

### 2.1. Participants and Screening Across Programme Stages

Across the three programme stages, age, sex assigned at birth and gender data were available for all 133 stage-level analysed records: 31 from Study 1, 32 from Study 2, and 70 from the Study 3 endpoint sample. Median age was 22 years (IQR 20–27; range 18–79). Sex assigned at birth was recorded as female for 87/133 records (65.4%) and male for 46/133 (34.6%); gender was recorded as female for 82/133 (61.7%), male for 46/133 (34.6%), and other/self-described for 5/133 (3.8%). Student status was not collected as a dedicated demographic variable and therefore could not be reliably reconstructed. Stage-specific demographic summaries are provided in the Supplementary Material.

Participants were aged 18 years or older, provided informed consent, and, before in-person exposure, completed the Sussex Strobe Safety Screening Questionnaire (4SQ; Schwartzman et al., 2025). Positive or uncertain safety responses resulted in exclusion or further eligibility review. Screening covered seizure/photosensitivity risk, recent traumatic brain injury, relevant neurological or ophthalmological conditions, severe psychiatric history, pregnancy, relevant medication exposure, substance-related risk, and marked sensitivity to bright or flickering light. Recruitment for Study 1 and Study 3 was conducted through the university and local community and required current self-reported non-seasonal depressive symptoms, including extended periods of low mood, and PHQ-9 score O5; formal MDD diagnosis was not required. The healthy Study 2 calibration sample instead used a STAI-Trait score <60 as an additional safety-screening criterion rather than as a depressive-symptom eligibility measure. The Study 1 and Study 3 samples are therefore adults reporting depressive symptoms rather than diagnostically confirmed MDD cohorts.

Across stages, exclusions included current antipsychotic treatment; use of supplements, herbs, or medicines for too short a period to establish familiarity; current or previous substance or drug misuse; current or previous psychosis, bipolar disorder, Parkinson’s disease, dementia, or Alzheimer’s disease; traumatic brain injury; and retinal blindness, cataracts, retinal disease, or glaucoma. Study 1 and Study 3 additionally excluded seasonal depressive symptoms and regular use of light therapy. Study 3 also excluded participation in Study 2 and plans to start or change treatment for depressive symptoms during the next four weeks.

During stimulation, adverse effects were actively monitored and a discomfort rating O7/10 triggered discontinuation of the relevant parameter set or session. Participants could stop or withdraw at any time. Mood and suicidality were monitored during visits and, in Study 3, between visits; worsening scores were flagged for review by the clinical or study lead under the monitoring procedure.

The monitoring documentation specified a PHQ-9 increase of ≥5 points from the first available observation and a two-point deterioration in positive or negative affect scores on the Scale of Positive and Negative Experience (SPANE-P or SPANE-N). It also specified a 12-point change in mood, pleasure, or suicidality ratings on the Maudsley 3-Item Visual Analogue Scale (M3VAS), transformed to a 0–100 scale. Meeting any of these thresholds was intended to prompt review by the clinical or study lead.

The complete screening workflow and supporting mood/suicidality monitoring documentation are provided in Supplementary Material Section S1.

### 2.2. Study 1: Parameter Optimisation

Study 1 participants attended one supervised laboratory visit. After baseline PHQ-9, Maudsley 3-Item Visual Analogue Scale (M3VAS) and Beck Depression Inventory-II (BDI-II) assessments, they completed 11 two-minute SLS sessions in a fixed staircase order. Sessions varied RX1 output, frequency, rapid frequency modulation, and duty cycle, with the final session combining these elements dynamically.

Frequency-specific Study 1 sessions sampled a broad active SLS range of approximately 3-15 Hz: fixed frequency trials progressed through 3, 5, 7 and 9 Hz, and subsequently 9, 11, 13 and 15 Hz, followed by trials examining progressively larger within-session frequency shifts. This range was used for exploratory parameter testing; it was not prespecified as a therapeutically optimal frequency band. Its purpose was to determine whether a broad range of SLS frequencies could be delivered tolerably in adults reporting depressive symptoms before construction of the longer Study 3 sequences.

After each session, participants completed the Visual Discomfort Questionnaire (VDQ; Vinkers et al., 2024), rated discomfort from 0 to 10, and stopped testing for that parameter if discomfort was O7; engagement, pleasure/enjoyment, discomfort, and arousal/sleepiness were rated, followed by post-sequence PHQ-9 and M3VAS-Change. The sequential Study 1 parameter manipulations are summarised in Table 1; additional parameter-selection details are provided in the Supplementary Material.

**Table 1.**
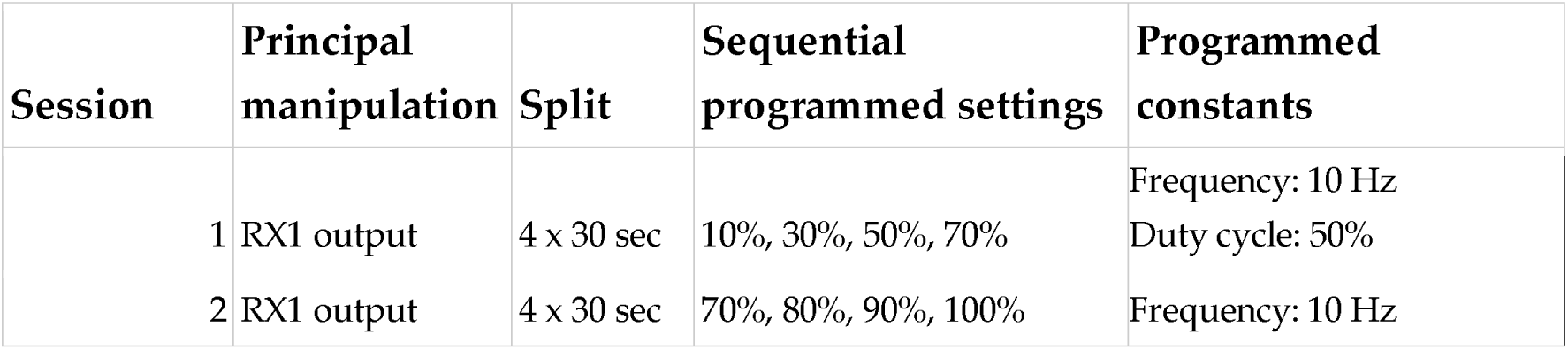
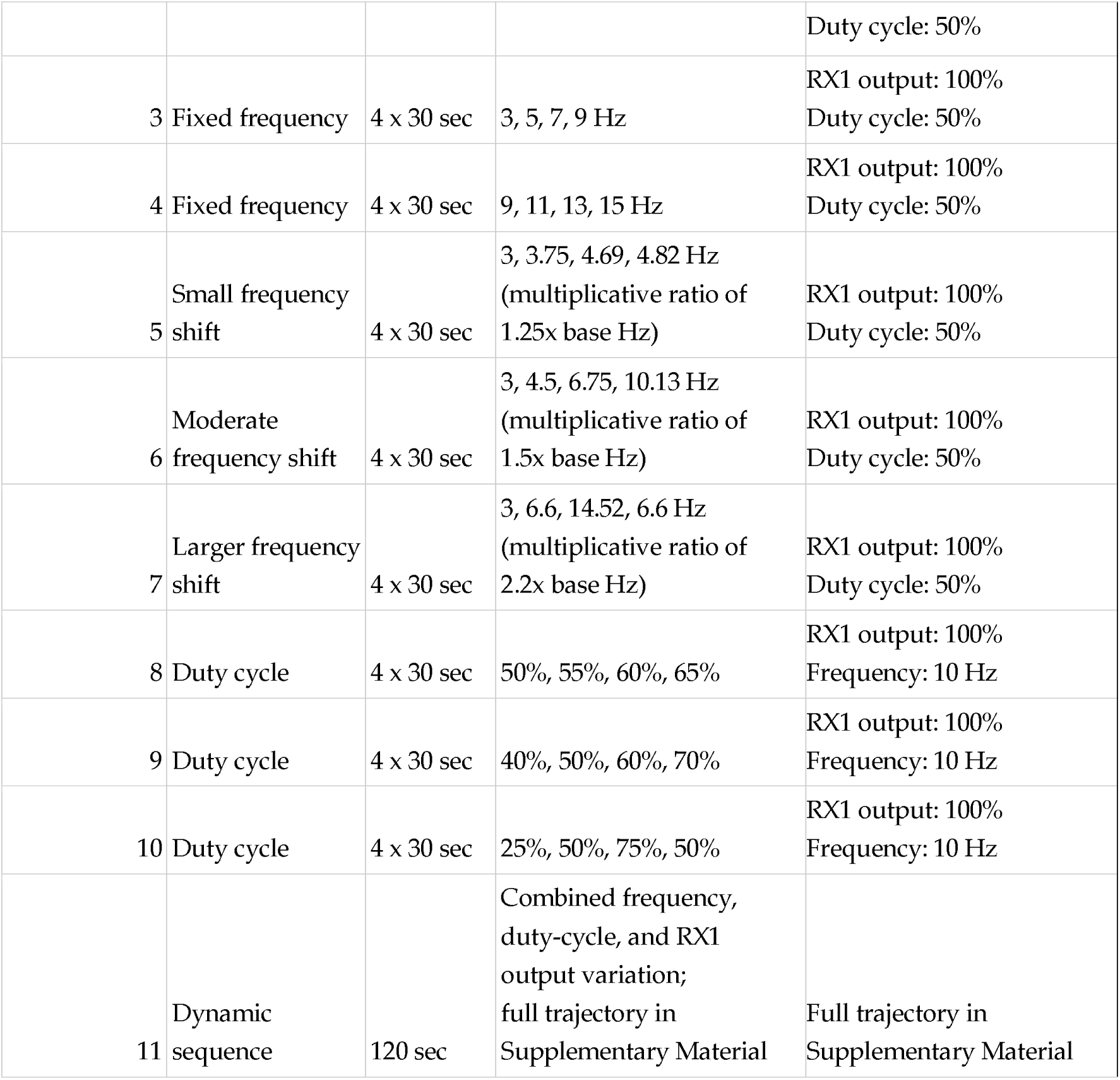
Sequential Study 1 SLS parameter manipulations.

### 2.3. SLS Configuration Development

The Study 3 stimulation protocol was constructed from parameter ranges that remained tolerable during Study 1. Study 1 did not determine how those parameters should be organised across a longer session. A reproducible structure inspired by classical sonata organisation was therefore used to arrange changes in SLS parameters over time. The resulting sequences contained a gradual opening, more variable and higher-intensity developmental portions, and a resolving final portion. This organisation is not proposed as a therapeutic mechanism, nor did the present study compare sonata-inspired organisation with alternative temporal structures.

Within these sequences, active stroboscopic stimulation varied approximately between 3.75 and 15 Hz, with programmed duty cycle varying from 25% to 75% and RX1 output from 0% to 100%. Separate high-rate 60-Hz wash-like phases and periods of darkness provided additional transitions and contrast. In the strobe-emphasis configuration, the wash-to-strobe programmed-output ratio was set at approximately 1:10 to produce a vivid but still tolerable visual experience. The wash-emphasis active comparator inverted this relation to approximately 10:1, reducing vivid stroboscopic effects while preserving session duration, the same temporal sequence structure, flicker cues, and the same broad programmed light-output profile. Because it retained visible flicker and a high-rate wash component, the control should be interpreted as an active, lower visual-phenomenology comparator rather than as an inert or perceptually equivalent control.

Each Study 3 configuration comprised 330 programmed time steps, with each step specifying start-to-end frequency, duty-cycle, and output values for two oscillator groups. The complete time-resolved specification is provided in the accompanying Supplementary Material Workbook (“Study 3 Parameters”).

### 2.4. Study 2: Comparator Calibration

Study 2 was conducted in healthy adults to calibrate the comparator rather than to test clinical efficacy or target-population generalisability. We therefore did not seek participants with depressive symptoms. Instead, participants without SLS contraindications and with STAI-Trait scores <60 completed a single-visit within-participant experiment comprising four supervised five-minute sessions in two randomised blocks, each containing a strobe-emphasis and wash-emphasis SLS sequence paired with frequency-tracking music. Musical accompaniment was also used in Study 3; Study 2 did not compare music with no music. The STAI threshold was used as part of SLS safety screening, not as a replacement for the PHQ-9 eligibility criterion used in Study 1 and Study 3.

After each session, participants completed the 6-Dimensional Visual Hallucination Questionnaire (6D-VHQ; Hewitt et al., 2026a) and immediate ratings of engagement, pleasure, arousal, and discomfort; after each block, they indicated which session produced the greater visual experience. The purpose of the bridge study was to test whether changing the balance between active flicker and washlight could alter visual experience in the same participants while preserving the wider stimulation context. It therefore provided evidence for developing the comparator, but did not establish that an equivalent difference in visual experience would generalise to adults reporting depressive symptoms. Study 3 then examined visual experience in this population directly. Full sequence and parameter details are provided in the Supplementary Material.

### 2.5. Study 3: Feasibility-Focused Randomised Trial

Study 3 was a parallel-group, allocation-concealed randomised feasibility trial allocating adults reporting depressive symptoms 1:1 to four weekly supervised sessions of approximately 31 minutes of either strobe-emphasis or wash-emphasis SLS. Both configurations involved active SLS. The Study 3 randomised contrast therefore estimates the difference in outcomes between two active SLS configurations delivered under matched supervision and audiovisual context; it does not estimate the effect of SLS relative to no stimulation, treatment as usual, constant light, or an inert sham. Allocation was stratified by psychoactive medication status and baseline PHQ-9 severity (scores of 5–16 vs 17–27), using randomly permuted blocks of four. Further allocation-sequence and concealment details are provided in the Supplementary Material. Configuration codes and template filenames were concealed from participants and testing researchers, as operational allocation-concealment measures; however, perceived allocation, credibility, and treatment guess were not measured, so masking success cannot be claimed. Additional exclusions included seasonal symptom patterns, regular use of light-based devices, and practical barriers to repeated attendance. Baseline assessments included BDI-II, Montgomery-Åsberg Depression Rating Scale, Self-Assessment version (MADRS-S), Beck Anxiety Inventory (BAI), Treatment Expectation Questionnaire (TEX-Q), and Stanford Expectations of Treatment Scale (SETS) (Svanborg & Åsberg, 2001; Beck et al., 1996; Beck et al., 1988; Shedden-Mora et al., 2023; Younger et al., 2012).

At each visit, participants reported interim treatment or symptom changes and completed the Frequency, Intensity, and Burden of Side Effects Rating (FIBSER) (Wisniewski et al., 2006). Before stimulation, PHQ-9, M3VAS/M3VAS-Change, and the Scale of Positive and Negative Experience (SPANE) assessed depressive symptoms, core symptoms/suicidality, and affect (Kroenke et al., 2001; Moulton et al., 2021; Diener et al., 2010). After the allocated SLS session, visually induced discomfort was assessed with the VDQ, symptoms and affect were reassessed using the PHQ-9, M3VAS-Change, and SPANE, and acute subjective experience was characterised using the 6D-VHQ, the 11-factor scoring of the 5D-ASC, and brief ratings of engagement, pleasure, discomfort, and drowsiness (Vinkers et al., 2024; Hewitt et al., 2026a; Studerus et al., 2010). SMS follow-ups after sessions 1–3 and a one-week post-treatment follow-up assessed short-term symptom persistence and affective change. Full scheduling details are provided in the Supplementary Material.

Depression-symptom assessments differed in timing and recall instruction across Study 3. Baseline PHQ-9 used its conventional two-week recall period; pre-session assessments for Sessions 2-4 referred to the preceding week; immediate post-session administrations referred specifically to the most recent SLS session; and the one-week follow-up again referred to the preceding week. These repeated values are therefore not interpreted as interchangeable administrations of the conventional PHQ-9. BDI-II was administered within the post-Session-4 questionnaire after completion of the fourth stimulation session.

### 2.6. Outcomes

#### 2.6.1. Study 1 Outcomes

Study 1 primary outcomes were safety and tolerability during brief supervised exposure. Safety was defined as the absence of severe adverse reactions directly attributable to SLS. Tolerability was assessed using 0-10 discomfort ratings, with success defined as the upper 80% confidence limit of the mean remaining <7; VDQ side-effect frequencies, discontinuation triggers, and individual intolerance events provided supporting data on tolerability. Feasibility was summarised descriptively through recruitment and completion. Exploratory outcomes included immediate PHQ-9, M3VAS-Change, and acute experience ratings.

#### 2.6.2. Study 3 Outcomes

Study 3 primary outcomes were safety, tolerability, and feasibility. Safety was summarised using the adverse-event and stopping-rule framework applied across the programme. Tolerability was indexed by 0-10 post-session discomfort ratings and side-effect summaries. Feasibility was assessed by recruiting 84 participants within 18 months, achieving ≥80% target recruitment as acceptable, and retaining ≥80% of participants who attended at least one session at treatment completion/post-session 4, with retention reported overall and by configuration because comparator retention is central to trial feasibility. Exploratory outcomes covered the symptom, affect, expectancy, and acute-experience measures listed above, plus later experience-outcome association analyses.

### 2.7. Analysis

Analyses were feasibility-first, descriptive, and estimation-focused. Binary and categorical outcomes were summarised as counts and proportions, and continuous outcomes summarised as means, standard deviations, and ranges. Clinical and experiential analyses were exploratory and hypothesis-generating rather than confirmatory. Study 2 was treated as a comparator-calibration study and is therefore not included in the Study 1/Study 3 primary-outcome analyses; full details are provided in the Supplementary Material.

#### 2.7.1 Study 1 Analyses

Study 1 safety was summarised descriptively. Tolerability was analysed session by session using 0–10 discomfort ratings, with means, standard deviations, and one-sided upper 80% confidence limits compared with the prespecified <7 criterion. VDQ symptom frequencies and acute experience ratings were tabulated descriptively; immediate PHQ-9 and M3VAS-Change were examined descriptively only.

#### 2.7.2. Study 3 Primary Outcome Analyses

Study 3 safety, tolerability and feasibility were analysed against programme-defined thresholds, with the source of each threshold documented in the Supplementary Material. Safety summaries included adverse reactions, serious adverse events, and safeguarding-triggered discontinuations. Tolerability summaries included discomfort ratings, side-effect frequencies, and tolerability-related discontinuations or incomplete completions, interpreted against the mean-discomfort criterion of <7 and alongside individual tolerability events. Feasibility summaries included recruitment, retention, discontinuation, and protocol adherence, interpreted against the progression criteria above and reported overall and by configuration.

#### 2.7.3. Study 3 Exploratory Clinical Analyses

Study 3 clinical analyses were exploratory, descriptive, and estimation-focused. Depressive symptom outcomes were summarised. PHQ-9 mean scores were summarised across baseline, repeated pre-session assessments, post-session 4, and, where available, one-week follow-up. BDI-II mean scores were summarised from baseline to post-session 4. Baseline-to-endpoint change was endpoint minus baseline, so negative values indicate symptom reduction for PHQ-9, BDI-II, MADRS-S, and BAI. Configuration-specific mean changes, strobe-emphasis-minus-wash-emphasis change contrasts, and baseline-adjusted endpoint contrasts for core endpoints were reported descriptively with 95% confidence intervals. These analyses were not confirmatory efficacy tests; retention imbalance and available-case missingness mean they should be read as feasibility-trial endpoint-planning estimates rather than evidence of efficacy.

Exploratory depressive-symptom changes were considered against minimal important difference (MID) benchmarks, including a 5-point PHQ-9 change and published BDI-II change thresholds of 17.5–20%. These were used as descriptive benchmarks rather than confirmatory decision rules (Button et al., 2015; Kounali et al., 2022). Response, remission, and short-term follow-up summaries were descriptive only.

#### 2.7.4. Study 3 Exploratory Experiential and Experience-Outcome Association Analyses

Session-specific 6D-VHQ, 11-ASC, and immediate ratings of engagement, pleasure, discomfort, and drowsiness were summarised descriptively by session and configuration. Exploratory experience-outcome association analyses were conducted at the participant level to examine whether acute subjective experience tracked subsequent depressive-symptom change.

Exploratory continuous within-configuration models examined whether z-scored total ASC predicted post-treatment BDI-II after adjustment for baseline BDI-II and baseline expectancy. These models were estimated separately within the strobe-emphasis and wash-emphasis configurations. All experience-outcome analyses were descriptive and hypothesis-generating. They were not powered tests of moderation, mediation, or mechanism.

#### 2.7.5. Missing Data

Participants who never commenced treatment were excluded from adherence summaries; those who later withdrew were retained in the feasibility outcomes. Outcome analyses used only available cases, with no imputation. Recruitment, attendance, adherence, retention, and questionnaire completion metrics were reconstructed from attendance logs, scheduled assessments, and survey files. Because endpoint availability differed by configuration, exploratory BDI-II contrasts should be interpreted cautiously as available-case estimates.

## 3. Results

### 3.1. Programme Flow

Across stages, 197 participants were screened for Study 1, 109 for Study 2, and 312 for Study 3; 31, 32, and 70 participants, respectively, contributed the primary analysable datasets shown in Figure 1. Across the full programme, 618 individuals were screened, and 133 contributed analysable data at least once. Figure 1 summarises programme flow, including Study 3 allocation and post-session-4 analysis availability by configuration.

**Figure 1.**
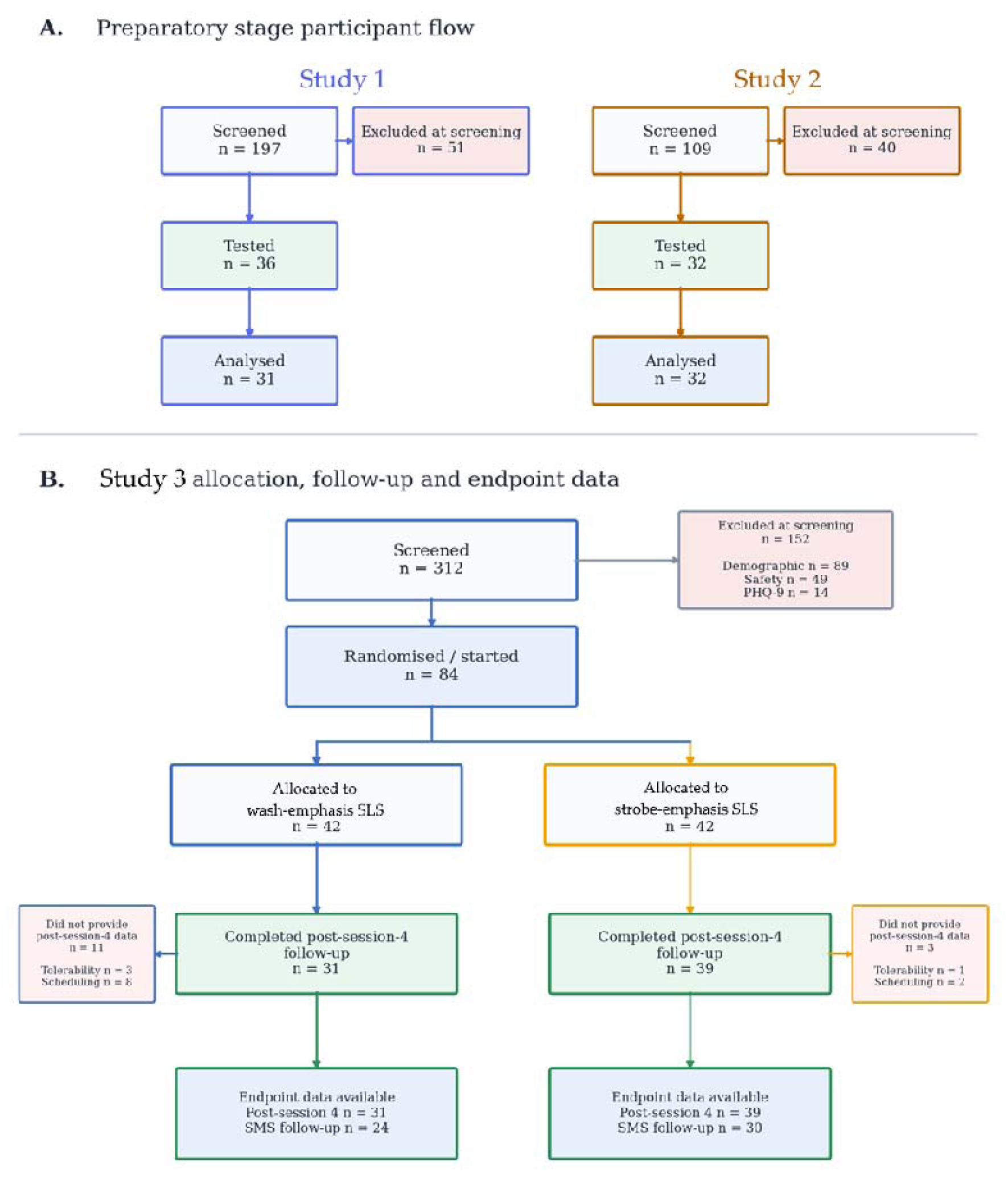
Programme flows across the staged SLS development programme. (A) Simplified flow for Study 1 and Study 2, showing the screened, tested, and analysed participants. Intermediate eligibility, booking, attendance, and non-attendance stages are omitted for cross-stage comparison. Study 1 screened 197 participants, of whom 148 passed screening, 36 were tested, and 31 were analysed; Study 2 screened 109, 32 were tested, and 32 were analysed. **(B)** Study 3 flow by trial configuration. Study 3 screened 312 participants and randomised 84; 42 were allocated to each configuration, with analysable post-session-4 data for 31 wash-emphasis SLS participants and 39 strobe-emphasis SLS participants; discontinuation/loss reasons are summarised in the Supplementary Material.

### 3.2. Study 1

#### 3.2.1. Safety and Tolerability of Brief SLS Exposure

Study 1 tested the safety and tolerability of brief SLS exposure in adults reporting depressive symptoms. Thirty-six participants were tested, and 31 (86%) contributed sufficient session-level discomfort data for analysis. Discomfort ratings remained very low across the 11-session staircase protocol: session-level mean discomfort ranged from 0.16 to 0.84 out of 10, with an average session-level mean of approximately 0.49/10. The highest one-sided upper 80% confidence limit was 1.13/10, observed in Session 1, remaining far below the prespecified discontinuation threshold of 7/10 (Figure 2). Thus, all Study 1 parameter classes met the predefined tolerability criterion, supporting their subsequent use in constructing the dynamic Study 3 stimulation sequences.

**Figure 2.**
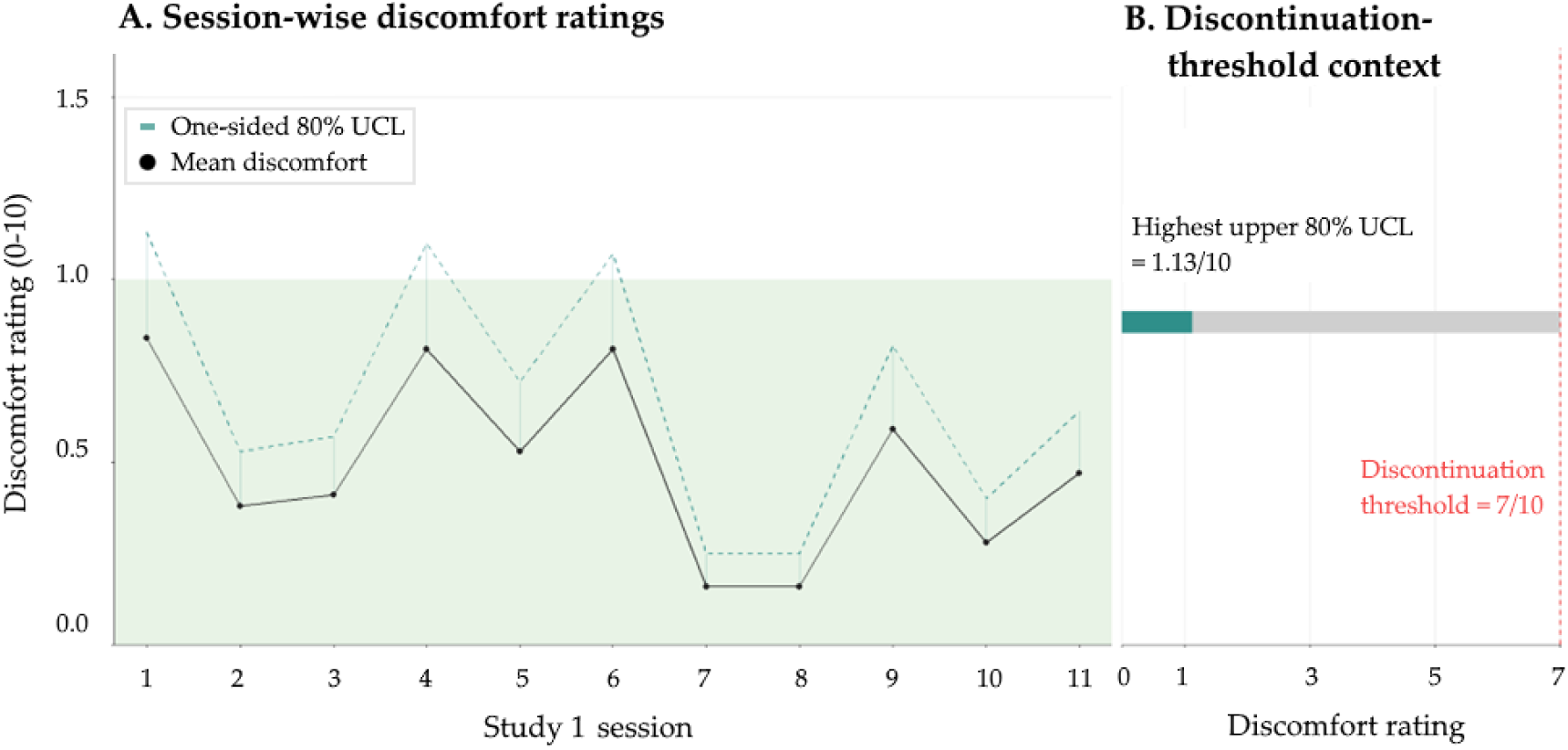
Study 1 session-wise discomfort ratings and discontinuation-threshold context. (A) Mean discomfort ratings across the 11-session Study 1 staircase protocol, with one-sided upper 80% confidence limits (UCL). Ratings used a 0–10 scale, where 0 indicated no discomfort and 10 indicated extreme discomfort. The shaded band marks ratings below 1/10 to show the observed low-discomfort range. **(B)** Threshold-context display showing the highest observed one-sided upper 80% confidence limit (1.13/10, Session 1) relative to the prespecified discontinuation threshold of 7/10.

#### 3.2.2. Parameter Selection, Acceptability, and Acute Symptomatology

Several Study 1 configurations combined acceptable discomfort with sufficiently vivid stroboscopic visual effects for later protocol development. Immediate post-session monitoring did not suggest short-term depressive symptom worsening, supporting progression to a repeated-session protocol. Full parameter-selection details and acute symptom-monitoring summaries are provided in the Supplementary Material.

### 3.3. Study 2: Comparator Calibration

Study 2 tested whether, in healthy adult participants, the low-phenomenology comparator reduced stroboscopic visual effects while preserving a shared session context. All 32 valid attendees were analysed using the 6D-VHQ to assess SLS-induced phenomenology.

Strobe-emphasis-minus-wash-emphasis paired mean differences were positive for five of six 6D-VHQ dimensions, with the largest differences for Geometric Content, Detail, and Vividness; Entropy showed no clear configuration difference. Strobe-emphasis SLS sessions were selected as producing the stronger visual experience in 46/64 binary rankings, compared with 18/64 rankings in the wash-emphasis sessions (one-sided binomial p < .001). Because each participant contributed two rankings, the binomial calculation is treated as a descriptive ranking-level check rather than a participant-level inferential test. The pattern was consistent with phenomenological separation, not masking success or comparator credibility, and supported carrying the low-phenomenology comparator forward for feasibility testing in Study 3 (Figure 3).

**Figure 3.**
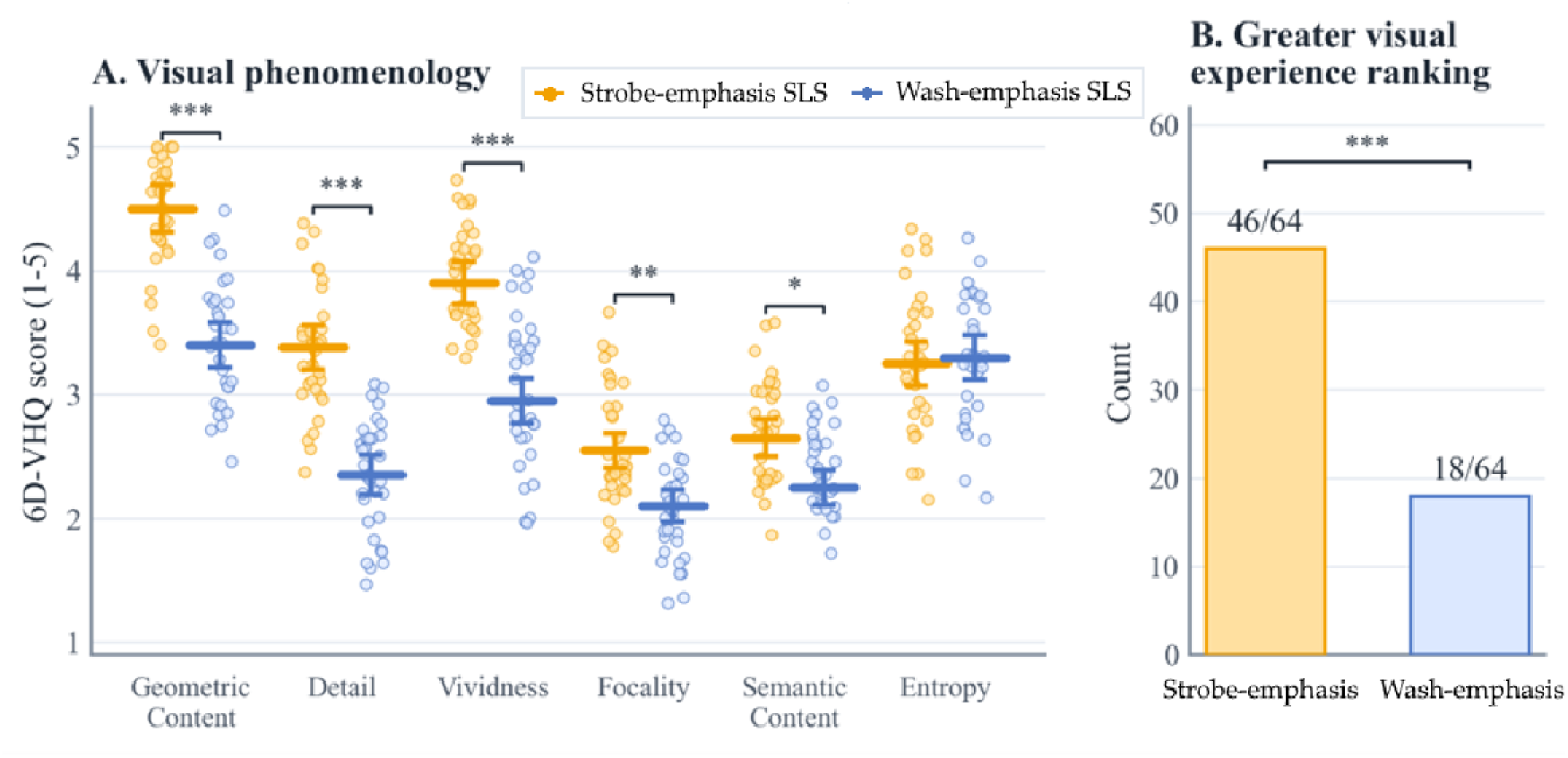
Study 2 support for the low-phenomenology comparator configuration. (A) 6D-VHQ visual phenomenology scores for wash-emphasis and strobe-emphasis SLS sessions. Horizontal lines show configuration means, error bars show standard errors, and points show individual session-level observations. Strobe-emphasis-minus-wash-emphasis paired mean differences were positive for Geometric Content, Detail, Vividness, Focality, and Semantic Content, whereas Entropy showed no clear difference. **(B)** Binary ranking counts show which session in each block produced the greater visual experience. Intervention sessions were selected more often. Panel A p-values are from exploratory repeated-measures models with participant-level random intercepts; asterisks denote *p < .05, **p < .01, ***p < .001. No formal multiplicity correction was applied. Panel B reports a descriptive, repeated-within-participant ranking count; the one-sided binomial value (p < .001) is a descriptive ranking-level statistic only, not a participant-level inferential test.

### 3.4. Study 3

#### 3.4.1. Repeated-Session Safety, Tolerability, and Feasibility

Study 3 tested full-length 31-minute SLS sessions after the brief 2-minute Study 1 exposure. Eighty-four participants were randomised. Session-by-session completion declined from 39/42 to 31/42 in wash-emphasis SLS and from 42/42 (100%) to 39/42 (93%) in strobe-emphasis SLS between Sessions 1 and 4. Endpoint availability differed substantially between randomised configurations. Post-Session-4 endpoint data were available for 39/42 participants assigned to strobe-emphasis SLS (92.9%) and 31/42 assigned to wash-emphasis SLS (73.8%). Available-case clinical contrasts are therefore based on samples retained at different rates after randomisation, but differential attrition alone does not establish bias. 14 of 84 randomised participants did not provide the post-Session-4 endpoint: 3 of 42 in strobe-emphasis SLS and 11 of 42 in wash-emphasis SLS. Of these, four withdrawals were attributed to discomfort (2 per configuration), both withdrawals attributed to perceived negative effects occurred in wash-emphasis SLS, and seven of eight scheduling/logistical losses occurred in wash-emphasis SLS. Mean Session-1 discomfort was also numerically higher under wash-emphasis SLS than strobe-emphasis SLS (0.72 vs 0.22 on the 0-10 scale), although both values were low in absolute terms. This descriptive difference does not establish that discomfort caused subsequent withdrawal. Taken together, the available data do not identify a single mechanism underlying the configuration difference in retention.

Baseline BDI-II, PHQ-9, and treatment expectancy were additionally summarised by endpoint-completion status within each configuration. Under strobe-emphasis SLS, endpoint completers and non-completers had mean baseline BDI-II scores (26.74, n = 39 vs 26.00, n = 3) and PHQ-9 scores (12.46, n = 39 vs 12.33, n = 3), while mean baseline expectancy was higher among completers (57.00 vs 39.67). Under wash-emphasis SLS, baseline data were available for all 31 endpoint completers and 8 of 11 non-completers; completers had higher baseline BDI-II scores (29.13 vs 24.12), slightly lower PHQ-9 scores (13.06 vs 14.50), and lower baseline expectancy (56.87 vs 71.12). These descriptive comparisons do not identify a consistent pattern of baseline severity or expectancy associated with endpoint completion, and the small non-completer samples limit conclusions about why attrition differed between configurations. Overall, Study 3 supports further development of a supervised intervention format, with retention in each configuration requiring its own prespecified progression rule in any future trial (Figure 4).

**Figure 4.**
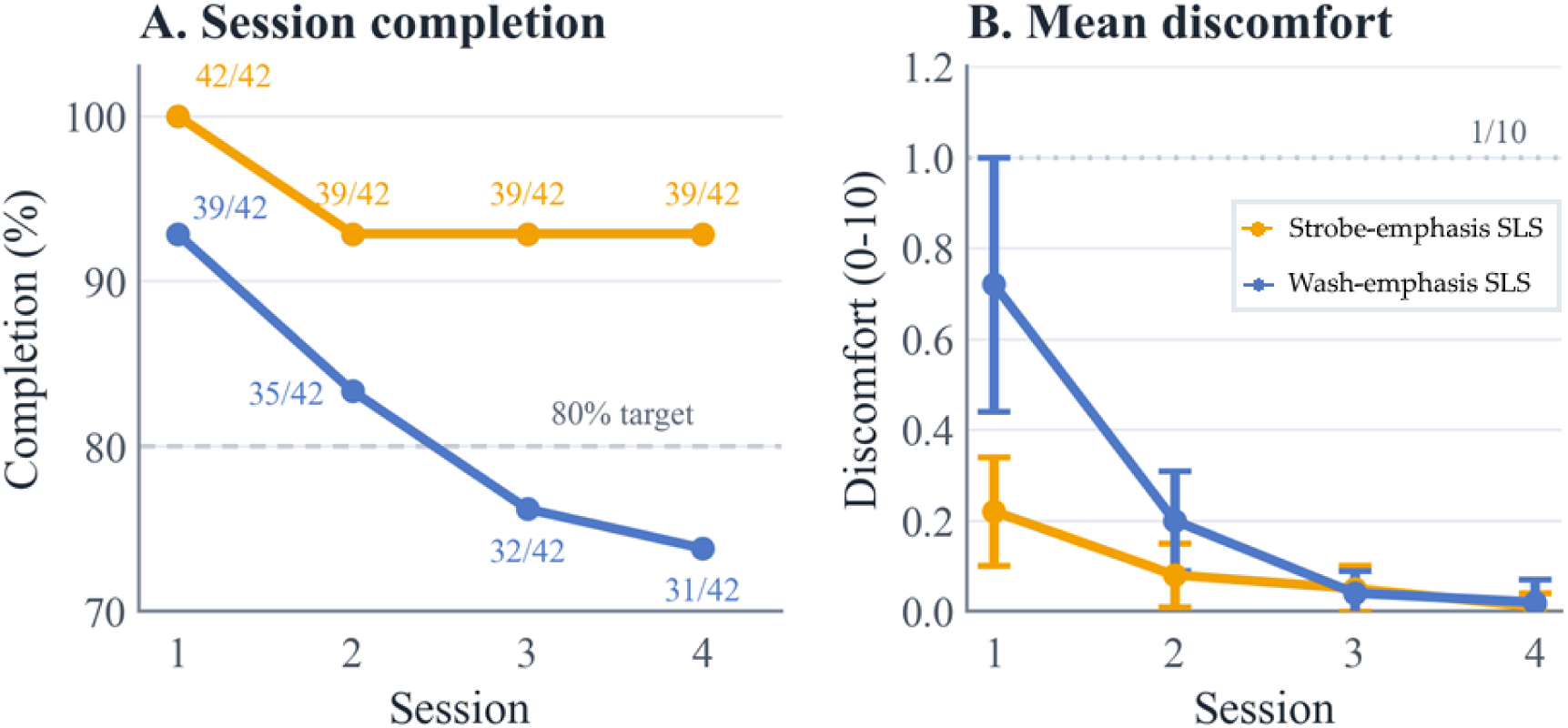
Study 3 feasibility and repeated-session discomfort ratings. (A) Session completion across the four-session Study 3 protocol, shown separately for wash-emphasis and strobe-emphasis configurations as percentages of randomised participants, with configuration denominators of 42 per group. Point labels show the number completed out of 42 randomised participants per configuration, and, when plotted on a 0-100% axis, the dashed horizontal line marks the 80% retention target. **(B)** Mean post-session discomfort ratings across sessions by study configuration. Ratings used a 0–10 scale, where 0 indicated no discomfort and 10 indicated extreme discomfort. Error bars show 95% confidence intervals. Discomfort is plotted on a zoomed 0-1.2 y-axis to show the floor-level observed range; the dotted line marks 1/10 for visual reference.

#### 3.4.2. Depressive Symptoms and Affective Outcomes

Study 3 depressive symptom and affective outcomes were exploratory and analysed descriptively using available cases with paired baseline-to-endpoint data. Change was defined as endpoint minus baseline. For BDI-II, negative values indicate lower endpoint symptom scores; repeated PHQ item sums require additional caution because recall instructions differed across assessment timepoints. On the paired endpoint sample, PHQ-9 mean change was −6.23 points in the wash-emphasis configuration (95% CI [−8.54, −3.91]; n = 31) and −8.26 points in the strobe-emphasis configuration (95% CI [−10.03, −6.48]; n = 39). The strobe-emphasis-minus-wash-emphasis difference in change was −2.03 points (95% CI [−4.85, 0.79], p = 0.155), and the baseline-adjusted endpoint contrast was −2.34 points (95% CI [−4.97, 0.28], p = 0.080). Mean PHQ-9 and BDI-II trajectories by configuration are shown in Figure 5.

**Figure 5.**
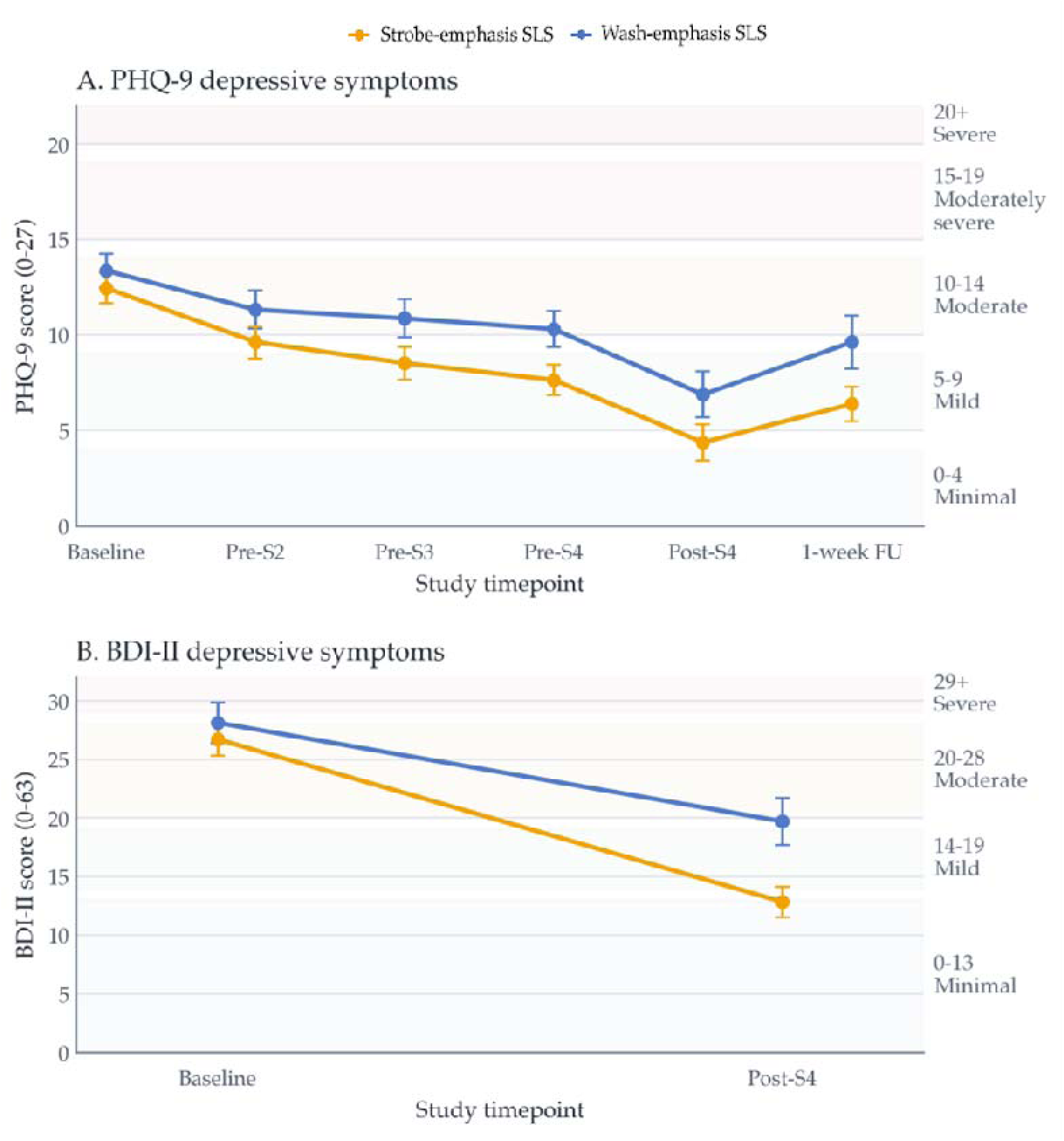
Study 3 exploratory depressive-symptom outcomes. (A) Mean PHQ-9 item-sum scores across repeated Study 3 assessments. Recall instructions differed across timepoints as described in Section 2.5, so these values should not be interpreted as interchangeable conventional PHQ-9 administrations. **(B)** Mean BDI-II scores from baseline to post-treatment endpoint. Lower scores indicate fewer symptoms. Error bars show 95% confidence intervals as plotted, and shaded bands indicate conventional symptom-severity ranges. Analyses were exploratory and available-case based. Negative baseline-to-endpoint differences indicate lower endpoint scores; PHQ item-sum differences are interpreted descriptively because recall instructions differed across timepoints. Strobe-emphasis-minus-wash-emphasis change was −2.03 points for PHQ-9 (95% CI [−4.85, 0.79], p = 0.155) and −4.57 points for BDI-II (95% CI [−10.08, 0.93], p = 0.102), and the baseline-adjusted endpoint contrast was −6.27 points (95% CI [−10.82, −1.72], p = 0.008). Estimates are descriptive and hypothesis-generating, and should not be interpreted as evidence of a configuration-specific clinical effect.

For BDI-II, mean change was −9.58 points in the wash-emphasis configuration (95% CI [−13.26, −5.90]; n = 31) and −14.15 points in the strobe-emphasis configuration (95% CI [−18.20, −10.11]; n = 39). The strobe-emphasis-minus-wash-emphasis difference in change was −4.57 points (95% CI [−10.08, 0.93], p = 0.102), and the baseline-adjusted endpoint contrast was −6.27 points (95% CI [−10.82, −1.72], p = 0.008). The unadjusted and baseline-adjusted estimates are presented together and neither is treated as confirmatory evidence. These available-case estimates are retained primarily to inform endpoint selection, variability, missingness, and sample-size planning for a subsequent trial.

For the modified PHQ item sums, at Pre-Session 4, the estimated strobe-emphasis-minus-wash-emphasis difference was −2.74 points (95% CI −5.32 to −0.15); immediately post-Session 4, it was −2.56 points (95% CI −5.51 to 0.40); and at one-week follow-up, among participants with available data, it was −3.24 points (95% CI - 6.29 to −0.20). Because these administrations used different recall instructions and follow-up availability was reduced, the three estimates should not be interpreted as repeated measurements of an identical conventional PHQ-9 construct.

#### 3.4.3. Acute Phenomenology and Active-Comparator Separation

Both Study 3 configurations produced substantial visual phenomenology (Figure 6). Across Sessions 1–4, wash-emphasis SLS means remained above the 1-point 6D-VHQ scale floor, including 4.18–4.36 for Geometric Content and 3.62–3.94 for Vividness. In mixed-effects models, pooled between-configuration differences (Δ = strobe-emphasis SLS − wash-emphasis SLS; corresponding to Intervention − Control under the registered analysis coding) averaged across sessions were small and imprecise: Geometric Content Δ = 0.21 (95% CI −0.07 to 0.50), Detail Δ = 0.17 (−0.20 to 0.54), Vividness Δ = 0.07 (−0.24 to 0.38), Focality Δ = 0.26 (−0.11 to 0.64), Semantic Content Δ = 0.16 (−0.26 to 0.58), and Entropy Δ ≈ 0.00 (−0.39 to 0.38). None of the confidence intervals for the pooled contrasts excluded zero.

**Figure 6.**
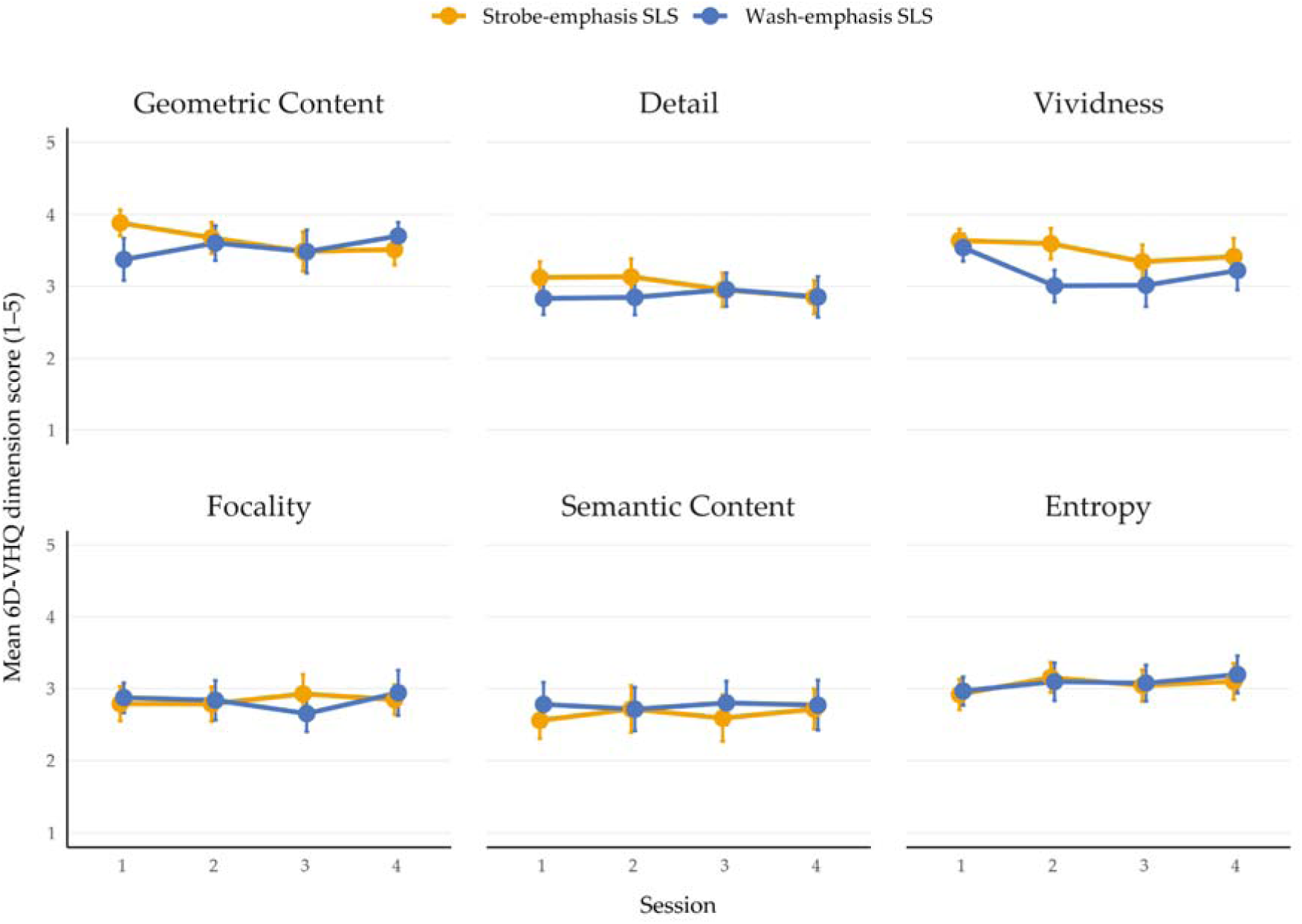
Study 3 acute visual phenomenology by SLS configuration and session. Six 6D-VHQ dimension means are shown on the original 1–5 scale for strobe-emphasis and wash-emphasis SLS. Points indicate observed configuration-specific means and error bars indicate 95% confidence intervals. Repeated-session summaries use available observations without imputation. Available sample sizes at Sessions 1–4 were 40, 39, 39 and 39 for strobe-emphasis SLS and 39, 35, 32 and 31 for wash-emphasis SLS, respectively. Mixed-effects analyses corresponding to these trajectories are reported in the accompanying text.

The direction of five of the six pooled effects was consistent with Study 2, but their magnitudes were smaller. The broad phenomenological separation observed during healthy within-participant calibration was therefore not reproduced at the group level in the parallel-group Study 3 target sample. Session-specific differences were observed for some dimensions, but these did not establish a broad and stable higher-versus-lower phenomenology distinction.

#### 3.4.4. Expectancy, Subjective Experience, and Exploratory Associations with Depressive Symptoms

Exploratory continuous within-configuration models examined whether acute total ASC score was associated with post-treatment BDI-II after adjustment for baseline BDI-II and baseline treatment expectancy. In the strobe-emphasis SLS configuration, each 1-SD higher total ASC score was associated with a −2.49-point difference in post-treatment BDI-II (95% CI −5.21 to 0.23, p = 0.071); in the wash-emphasis SLS configuration, the corresponding estimate was −1.75 points (95% CI −6.08 to 2.59, p = 0.415). Both confidence intervals included zero, and the models did not establish an association in either configuration.

## 4. Discussion

This staged developmental programme evaluated SLS in adults reporting depressive symptoms, prioritising safety, tolerability, feasibility, and active-comparator development rather than definitive efficacy testing. Study 1 identified a low-discomfort parameter space, with mean discomfort below 1/10 and all session-level upper 80% confidence limits well below the prespecified threshold. Study 2 supported a low-phenomenology comparator configuration that reduced subjective visual effects while preserving the session context, but it did not test masking or comparator credibility. Study 3 showed that four weekly supervised sessions could be delivered with recruitment to target, acceptable completion, low discomfort, and no serious SLS-attributable adverse events. Exploratory symptom outcomes support further testing but remain hypothesis-generating, with imprecise between-configuration estimates. Together, these findings reduce key uncertainties around repeated supervised SLS delivery and identify comparator retention, masking, and endpoint precision as priorities for a larger trial.

### 4.1. Safety, Tolerability, Feasibility, and Comparator Development

The safety and tolerability of SLS are central to its development, as SLS uses rhythmic visual stimulation to alter perception and subjective experience. The main medical risk is photosensitive convulsive seizure, which appears rare in the general population and concentrated in a susceptible photosensitive subgroup rather than in all people with epilepsy (Schwartzman et al., 2025). The Dreamachine installation adds non-clinical operational evidence from more than 40,000 people who experienced supervised closed-eye SLS in an immersive audiovisual context (Dreamachine, n.d.; Beauté et al., 2026). However, that public deployment does not establish safety or tolerability in people reporting depressive symptoms. Clinical depression samples may differ in medication use, sleep disturbance, anxiety, and sensory sensitivity. Direct evaluation in this group was therefore necessary.

Study 1 provided this first test. No severe adverse reactions occurred, mean discomfort was 0.49/10, and all session-specific upper 80% confidence limits stayed below the prespecified 7/10 threshold. Minor side effects were generally mild and transient; the most common examples were mild headache (18), pressure behind the eyes (18), urge to blink (16), and participant-defined symptoms (16), supporting the safety and tolerability of brief supervised SLS within the tested parameter range. The ∼15-Hz upper boundary should not be interpreted as an intrinsic safety threshold; safety in the present project also depended on structured contraindication screening, controlled exposure conditions, and close supervision.

Study 3 extended Study 1 by testing whether participants with depressive symptoms could tolerate longer 31-minute SLS sessions and complete four weekly sessions in a format closer to a plausible supervised intervention. The protocol reached the planned randomisation target of 84 participants; in-person session delivery was near-complete after Session 1 (Sessions 2-4: 74/74, 71/71 and 70/70), while overall scheduled-event completion across all channels was 87.6% (1,174/1,340). Mean discomfort stayed low throughout, and declined towards floor levels in later sessions. Fourteen participants discontinued or did not complete endpoint assessment (11 wash-emphasis, 3 strobe-emphasis); of these, four were discomfort-related (2 per configuration), two reflected perceived negative effects (both wash-emphasis), and eight were scheduling- or logistically-related (7 wash-emphasis, 1 strobe-emphasis). No serious SLS-attributable adverse events were identified. These outcomes map onto recommended progression-criteria domains for pilot and feasibility trials, in which prespecified indicators such as recruitment, retention, adherence, data completeness, acceptability, and tolerability are used to judge whether, and how, a future definitive trial should proceed (Mellor et al., 2021; Mellor et al., 2023). Together, these findings show that repeated supervised exposure met key recruitment, overall retention, tolerability, and safety criteria, while also identifying comparator-configuration retention as a design risk for the next trial. The available data do not identify a single mechanism underlying the configuration difference in retention; early-session discomfort, scheduling burden, symptom fluctuation, expectancy, and other practical factors may all be relevant, but none can be assigned a causal role from the present data. Future trials should measure engagement, credibility, and reasons for discontinuation as the trial proceeds.

No serious SLS-attributable adverse events were recorded in the randomised Study 3 cohort (n = 84). These observations constrain rather than exclude rare serious harms, including photosensitive convulsive seizures. With zero observed events and assuming complete safety follow-up for all 84 participants over the study period, the exact one-sided 95% upper confidence bound on the proportion experiencing such an event during the study is approximately 3.5%. Moreover, 49 of the 312 people screened for Study 3 were excluded on safety grounds. Safety conclusions should therefore be restricted to comparably screened and supervised populations and should not be generalised to individuals excluded because of elevated photosensitivity risk or other contraindications.

The active comparator is an important methodological output, but the Study 3 data qualify its interpretation. Study 2 demonstrated clear within-participant phenomenological separation in healthy volunteers; however, the same broad separation was not reproduced at the group level in Study 3. Instead, both Study 3 configurations produced substantial visual phenomenology, indicating that the randomised trial compared two active SLS configurations rather than comparing one active SLS configuration against a control that produced little visual experience. Some dimension- and session-specific differences remained, but the magnitude and temporal stability of the intended distinction were weaker in Study 3.

An active comparator could yield a smaller symptom contrast than a no-stimulation comparator if both SLS configurations have therapeutic effects. This would not, in itself, bias the estimate of their difference. Visual phenomenology alone does not establish therapeutic activity or a shared antidepressant mechanism.

The weaker observed separation in Study 3 might reflect, at least in part, differences in how phenomenology was assessed. Study 2 participants experienced both configurations and therefore had a direct basis for comparison, whereas Study 3 participants experienced only their allocated configuration. The present data therefore cannot distinguish genuine phenomenological overlap from possible differences in how participants used the rating scales. Future comparator validation should therefore be conducted directly in the target population with prespecified assessments of masking and credibility, and shared instructions for rating visual experience that do not reveal allocation.

### 4.2. Exploratory Clinical and Experiential Findings

Study 3 depressive-symptom outcomes were exploratory estimates derived from available cases. Their interpretation is constrained by differential endpoint availability and by differences in assessment timing and recall frame. In particular, immediate post-session PHQ responses used a session-specific recall instruction and should not be interpreted as conventional longitudinal PHQ-9 symptom change; BDI-II was administered after Session 4, and responses may therefore also have been influenced by how participants felt after stimulation.

Clinical outcomes should be interpreted within the feasibility framework of Study 3. Both SLS configurations showed descriptive reductions in symptom scores, while most between-configuration confidence intervals remained compatible with no difference. The baseline-adjusted BDI-II endpoint contrast excluded zero, but the corresponding unadjusted change contrast did not; moreover, endpoint availability differed markedly between configurations, and BDI-II was administered after Session 4. No tipping-point analysis is presented, so sensitivity to outcomes among participants with missing endpoints remains unquantified. Both estimates remain exploratory; the difference in whether their confidence intervals exclude zero does not itself establish a disagreement between them. The adjusted estimate does not, on its own, establish a differential clinical effect between configurations.

The symptom data are most useful for planning a subsequent trial; they provide estimates of outcome variability, the extent of missing data, and endpoint availability while identifying assessment timing as an important design parameter. A future trial should prespecify a validated symptom recall period and separate its primary depressive-symptom endpoint from the immediate stimulation period.

The unadjusted and baseline-adjusted estimates are presented together, and the adjusted BDI-II estimate is not given greater evidential weight solely because its confidence interval excludes zero. These results primarily provide information on variability and missingness to inform endpoint selection and sample-size planning for a subsequent trial.

Minimal important difference (MID) benchmarks help contextualise clinical relevance in an underpowered feasibility study (Kroenke et al., 2001; Whisman et al., 2000; Button et al., 2015; Kounali et al., 2022). Mean PHQ-9 change exceeded the 5-point threshold used in the programme’s descriptive MID summaries, although its confidence interval crossed below that benchmark. Mean BDI-II reduction more clearly exceeded the pragmatic 17.5% to 20% MID range (Button et al., 2015; Kounali et al., 2022). These within-configuration comparisons are descriptive only and should not be treated as evidence of a between-configuration treatment effect. BDI-II may merit endpoint-selection discussion because it is a longer, more detailed depression measure than the PHQ-9, but that decision should be based on the next trial’s clinical objective, burden, missingness profile, and prespecified statistical plan.

Similarly, adjusted continuous experience-outcome models did not establish an association between acute total ASC and post-treatment BDI-II in either configuration. Acute experience and depressive symptoms were both self-reported, and adjustment for baseline expectancy does not rule out confounding by expectations or biases shared by both measures. Total ASC remains a theoretically motivated candidate predictor for future adequately powered investigation, but the present data do not establish a dose-response, moderation, mediation, or mechanistic relationship.

The clinical rationale is therefore modest: SLS is sufficiently developed to warrant further evaluation as a distinct, experience-centred, standardisable, and experimentally controllable intervention, but the present evidence supports trial planning rather than a treatment claim.

### 4.3. Strengths and Limitations

Strengths include the staged design, explicit safety thresholds, low-phenomenology comparator development, and randomised repeated-session feasibility testing with operational allocation concealment. The programme generated practical translational outputs: tolerable parameter settings, a deliverable session structure, and a comparator rationale that can be carried forward into a later trial.

Limitations include the symptom-defined rather than diagnostically confirmed depression sample; single-site early-phase design; absence of dedicated student-status data, limiting assessment of sample composition and generalisability; insufficient power to test efficacy; short follow-up; visual phenomenology in the wash-emphasis SLS configuration; lower endpoint retention under wash-emphasis SLS; no assessment of masking success, credibility, or treatment guesses; unmeasured individual-difference variables, such as trait suggestibility or phenomenological control (Lush et al., 2020); and exploratory analyses of the relationship between acute experience and symptoms. A subsequent trial should assess masking, credibility, discontinuation reasons, and these individual-difference variables prospectively. Candidate-predictor analyses were further limited by small tertile sizes, baseline severity differences, and available-case missingness; future trials should prespecify experience-outcome models and distinguish prognostic, moderation, and mediation hypotheses. These limitations identify the design features that any subsequent study should strengthen.

### 4.4. Implications for the Next Phase

The next phase should use a pre-registered, locked protocol in a diagnostically defined depression sample, with prospective assessment of masking, expectancy, credibility, phenomenological-control traits, and durability. The present work supplies the operational ingredients for that step: tolerable parameter settings, a deliverable four-session structure, a low-phenomenology comparator requiring further credibility and retention testing, and a workable supervised monitoring workflow. The next trial should prespecify one primary clinical endpoint and timepoint, refine the control, report MID-anchored confidence intervals, and define red-amber-green progression criteria for safety, configuration-specific retention, missingness, and feasibility. If the experience-centred rationale remains central, acute experiential targets and masking criteria should be defined in advance.

## 5. Conclusion

In a screened and supervised population, SLS was feasible to deliver over repeated sessions with low discomfort. These findings support further controlled clinical development but do not exclude rare serious harms. The staged programme also identified important design limitations: retention was lower under wash-emphasis SLS, both Study 3 SLS configurations produced substantial visual phenomenology, and the broad separation in experience observed during within-participant calibration in healthy volunteers was not reproduced at the group level in the target population. Exploratory symptom outcomes can inform the choice of endpoints for future trials, but do not establish efficacy. The next step is therefore a feasibility trial in a diagnostically defined population, overseen by a clinical trials unit. It should have a locked protocol, a prespecified clinical endpoint and assessment timepoint, planned masking and credibility assessment, and explicit progression criteria for safety, retention in each configuration, missing data, and comparator performance.

## Data Availability

The WP1 and WP2 studies were registered as ISRCTN82430224 and ISRCTN13880276. In line with the registrations and the MRC DPGF data-management plan, anonymised analysis datasets supporting the reported results, data dictionaries, analysis code, and associated reproducibility materials are available at https://github.com/dannynacker/SLSD/tree/data. Data sharing remains subject to participant consent, ethics approval, UK GDPR, University of Sussex governance and, where required, a data-sharing agreement. Identifiable data, linkage keys, administrative records, precise attendance dates, and free-text safety or adverse-event material are not publicly shared.

## Acknowledgements

The authors are grateful to Cecelia Schwartzman and Lionel Barnett for their support and helpful discussions during the development of this manuscript.

## Author Contributions / CRediT

Conceptualization: D.J.S., A.K.S., J.M.S., D.N.

Methodology: D.J.S., D.N., J.M.S., A.K.S.

Investigation: D.N., L.K. and D.J.S.

Formal analysis: D.N., D.J.S.

Data curation: D.N., L.K. and D.J.S.

Software and visualisation: D.N. and D.J.S.

Resources: G.L., J.S., D.P., E.W., A.K.S. and D.J.S.

Supervision: D.J.S., A.K.S., J.M.S., J.W.S.

Project administration: D.N., L.K. and D.J.S.

Funding acquisition: D.J.S., J.M.S., S.B., W.W., A.K.S.

Writing – original draft: D.J.S., D.N. and R.C.-C.

Writing – review and editing: D.N., L.K., A.K.S., J.M.S., G.L., J.S., J.W.S., S.B., C.I.J., W.W., F.M., D.P., E.W. and D.J.S.

## Funding

D.J.S. is supported by a Medical Research Council Grant UKRI083. A.K.S. is supported by the European Research Council (ERC) Advanced Investigator Grant 101019254, under the European Union’s Horizon 2020 programme. J.W.S. is based at the NIHR University College London Hospitals Biomedical Research Centre, which receives funding from the UK Department of Health. He receives research support from the Dr Marvin Weil Epilepsy Research Fund, the Academy of Medical Sciences, and the National Institute for Health and Care Research.

## Declaration of Competing Interest

G.L. is affiliated with Audyssey, and J.S. is affiliated with roXiva Innovations Ltd. roXiva Innovations Ltd provided the roXiva RX1 stroboscope used in this study but had no role in the study design, data collection, data analysis, interpretation of the findings, or preparation of this manuscript. Beyond the relationships with Audyssey and roXiva Innovations Ltd disclosed above, no author reports any additional financial or proprietary interest, including employment, equity, patents, licensing, or paid consultancy, relating to an SLS device or manufacturer.

## Data Availability Statement

The Study 1 and Study 3 studies were registered as ISRCTN82430224 and ISRCTN13880276. In line with the registrations and the MRC DPGF data-management plan, anonymised analysis datasets supporting the reported results, data dictionaries, analysis code, and associated reproducibility materials are available at https://github.com/dannynacker/SLSD/tree/data. Data sharing remains subject to participant consent, ethics approval, UK GDPR, University of Sussex governance and, where required, a data-sharing agreement. Identifiable data, linkage keys, administrative records, precise attendance dates, and free-text safety or adverse-event material are not publicly shared.

## Declaration of generative AI and AI-assisted technologies in the manuscript preparation process

During the preparation of this manuscript, the authors used OpenAI’s ChatGPT/Codex to support language editing, manuscript organisation and journal-submission checks. After using these tools, the authors reviewed and edited the content and take full responsibility for the content of the published article.

## Notes

### Clinical Trial

ISRCTN13880276 and ISRCTN82430224

### Author Declarations

The study was conducted at the Sussex Centre for Consciousness Science, University of Sussex, United Kingdom. Ethical approval was granted by the University of Sussex Sciences and Technology Cross-Schools Research Ethics Committee, reference ER/LK344/4. All participants provided informed consent before taking part.

### Summary of Updates

This revision restores Robert Chis-Ciure to the author list and updates the author contributions statement. The scientific content is unchanged.

